# Do people with disabilities have the same level of HIV knowledge and access to testing? Evidence from 513,252 people across 37 Multiple Indicator Cluster Surveys

**DOI:** 10.1101/2023.07.18.23292845

**Authors:** Sara Rotenberg, Shanquan Chen, Jill Hanass-Hancock, Calum Davey, Lena Morgon Banks, Hannah Kuper

## Abstract

**Background:** Disability and HIV are intricately linked, as people with disabilities are at higher risk of contracting HIV and HIV can lead to impairments and disability. Despite this well-established relationship, there remains limited internationally comparable evidence on HIV knowledge and access to testing for people with disabilities.

**Methods and Findings:** We used cross-sectional data from 37 Multiple Indicator Cluster Surveys. 513,252 people were eligible for inclusion, including 24,695 (4.8%) people with disabilities. We examined risk ratios and their 95% confidence intervals for key indicators on HIV knowledge and access to testing for people with disabilities by sex and country. We also conducted a meta-analysis to get a pooled estimate for each sex and indicator. Men and women with disabilities were less likely to have comprehensive knowledge about HIV prevention (aRR: 0.74 [0.67, 0.81] and 0.75 [0.69, 0.83], respectively) and to know of a place to be tested for HIV (aRR: 0.95 [0.92, 0.99] and 0.94 [0.92, 0.97], respectively) compared to men and women without disabilities. Women with disabilities were also less likely to know how to prevent mother-to-child transmission (aRR: 0.87 [0.81, 0.93]) and ever have been tested for HIV (aRR: 0.90 [0.85, 0.94]), while men with disabilities showed some evidence of relative inequities for these indictors. There was also some evidence women with disabilities were less likely to be tested for HIV in the past year.

**Conclusion:** Men and women with disabilities face inequities in HIV knowledge and access to testing, particularly for women with disabilities. Governments must include people with disabilities in HIV programs by improving accessibility and increasing disability-inclusion in each health system building block.

## Introduction

Globally, there are 1.3 billion people with disabilities.[1] Disability and HIV are intricately linked, as research suggests that people with disabilities are at higher risk of contracting HIV (e.g. due to poverty, exclusion, and discrimination) and HIV can lead to impairments and disability (e.g. due to the direct effect of the virus, opportunistic infections or exclusion, and discrimination).[2–4] While HIV control efforts have centered around expanding access to prevention, testing, and treatment, these programs often fail to consider provision of accommodation for people with disabilities and those people living with HIV who develop disabling conditions.[4, 5] People with disabilities face widespread barriers in accessing health care, but these are amplified for HIV care by a lack of knowledge and accessible information about HIV/AIDS and access to sexuality education; cultural beliefs around disability and HIV/AIDS; lack of affordable, accessible, acceptable, and quality HIV care; and health workers’ beliefs that people with disabilities are asexual.[6–8]

Data on HIV knowledge among people with disabilities varies across surveys with different methods and different countries. While early disability focused studies suggested that persons with disabilities had lower knowledge about HIV, [9–13] recent studies using population-based surveys show more diverse result. For instance the 2017 HIV Impact Survey in Tanzania suggest that women with disabilities be more likely to know their HIV status.[14] In South Africa the 2012 National HIV Prevalence, Incidence and Behavior surveys found that people with disabilities had less knowledge about HIV and were less likely to find testing sites,[15] while the analysis of the 2011 Demographic Health Survey in Uganda showed equal knowledge of transmission for delivery and breastfeeding, but wide gaps in knowledge about other infection or transmission risk and misconceptions.[16]

Beyond this data, indicators on HIV prevention, testing, and treatment are rarely disaggregated by disability status in large-scale household surveys and country surveillance data and there is no comparable analysis of disability and HIV data across countries available so far. Hence, the UNICEF-supported Multiple Indicator Cluster Surveys (MICS) conducted across a large number of Low and Middle Income Countries (LMICs) is an opportunity to assess the HIV-knowledge and testing practices among people with disabilities. While the Disability Data Initiative reports suggest some differences in having heard of HIV or having ever been tested for HIV among women with disabilities compared to women with moderate or no disabilities, these reports only discuss descriptive statistics for women.[17] However, more country-specific and sex-disaggregated analysis is needed to further understand inequities.

This paper presents sex-disaggregated, internationally comparable evidence on HIV knowledge and testing among adults 15-49 from the UNICEF-supported Multiple Indicator Cluster Surveys (MICS) conducted in 37 LMICs. The aim is to compare in access to knowledge about prevention between people with disabilities and those without disabilities in access to knowledge about prevention and testing for HIV by disability status. Efforts to improve access to knowledge, testing, and treatment are central to UNAIDS 95-95-95 targets by 2030 and so these data provide evidence on how these efforts are reaching people with disabilities in HIV programmes.[3]

## Materials and Methods

The UNICEF-supported Multiple Indicator Cluster Surveys (MICS) are cross-sectional, population-based survey conducted in low- and middle-Income Countries. The MICS use a multi-stage sampling approach to sample clusters of households to generate nationally-representative data on indicators for tracking the Sustainable Development Goals, health, and development.[18, 19] The current analyses focus on the 37 countries where HIV and disability data were available for women (and a subset of 29 countries had men‘s disability and HIV data). Countries were geographically diverse, with 12 in sub-Saharan Africa, 7 in East and Central Asia, 6 in Latin and Central America, 6 in East Asia and Pacific, 4 in Middle East and North Africa, and 2 in South Asia.

Trained data collectors conducted household interviews with individual adults aged 15-49 living in the randomly selected households. All men and women aged 18-49 were eligible, while participants aged 15-17 may have been one of the children aged 5-17 randomly selected from the household. Data were collected for both individual women and men, where countries have opted-in to including the individual men’s questionnaire. Questions were standardized across countries, allowing comparisons across all the countries in which the sixth round of the MICS has been completed. We selected any country’s complete MICS survey that had anonymized individual data on all variables of interest and were publicly available as of March 2023, though data were collected between 2017-2021.

### Disability

Disability was measured in the child and adult functioning modules for adults 15-17 and 18-49, respectively. Both modules use the Washington Group Questions that assess the participants’ impairments based on their self-reported level of functional difficulty for each of the domains (**Table 1**). We defined disability as the highest two thresholds of impairment, including only those who answered ‘cannot do at all’ or a ‘lot of difficulty’ in at least one functional domain as disabled. However, it does mean our comparison group includes individuals who have some functional difficulty in one or more functional domains. This threshold of disability was selected in accordance with the Washington Group syntax, so that the indicators aligned with the UNICEF reports, and to preserve the specificity of the disability measure. Individuals without fully completed functioning modules were excluded from analysis, unless they had met the threshold for disability in one or more domains, since the missing data would not have impacted their disability status.

**Table 1:** Definitions of HIV anddisability indicators.

| **Indicators** | **MICS Indicator** | **Definition** |
| --- | --- | --- |
| Comprehensive knowledge about HIV prevention | TM.29 | % of people who know of the two ways of HIV prevention (having only one faithful uninfected partner and using a condom every time), who know that a healthy-looking person can be HIV-positive, and who reject the country-specific two most common misconceptions about HIV transmission. |
| Knowledge of mother-to-child-transmission (MTCT) | TM.30 | % of people who can identify that HIV can be transmitted from mother to child (during pregnancy, during delivery, and by breastfeeding). |
| People who know where to get tested for HIV | TM.32 | % of people who know where they can get tested for HIV. |
| People who have ever been tested for HIV and know the results | TM.33 | % of people who report ever being tested for HIV and know the results of the most recent test. |
| People who have been tested for HIV in the last year and know the results | - | % of people who report being tested for HIV in the last 12 months and know the results of the most recent test. |
| Disability | - | % of adults aged 18-49 with functional difficulty as assessed by the Washington Group Short Set ("cannot do at all" or "a lot of difficulty" in at least one of the following domains: 1) seeing, 2) hearing, 3) walking, 4) remembering/concentrating, 5) self-care, 6) communication) and adults 15-17 with functional difficulty as assessed by the Child Functioning Module ("cannot do at all" or "a lot of difficulty" in at least one of the following domains: 1) seeing, 2) hearing, 3) walking, 4) remembering 5) concentrating, 6) self-care, 7) making friends, 8) controlling behaviour, 9) accepting change, 10) learning, 11) communication or "daily" in either 12) anxiety, and 13) depression. |

### Outcomes and co-variates

Outcomes were related to HIV knowledge and testing behaviour, assessed through five questions (**Table 1**). We used the standard MICS definitions to calculate each outcome. Responses were reported by the individual participants, and those unable to participate were recorded as ‘incapacitated’[20]. Not all countries included had all HIV indicators or data on both sexes, which is why we did not conduct a ‘overall’ result that included both sexes. We adjusted the analysis by age (years), wealth quintile, and residence area (urban vs rural)

### Statistical Analysis

All analyses were completed using R 4.2.2. We described all outcomes, exposures, and covariates by country and overall. Continuous data were reported as mean (standard deviation [SD]), and categorical data were reported as numbers (percentage).

To estimate the relative inequities of each outcome between people with and without disabilities, we first modelled the probability of each outcome by sex and by country, using a modified Poisson model.[21] Results were reported as (adjusted) risk ratio (RR or aRR) and its 95% confidence interval (CI). The complex survey design and MICS sample weights were also accounted for using the ‘survey’ package in R.[22] We then pooled the country-specific estimations by meta-analysis with the inverted standard error as the weight. The heterogeneity of estimates across countries was assessed by Cochran’s Q test.[23] For the presence of significant heterogeneity (p < 0.1), a random-effect meta-analysis was performed to pool the estimates and for those where p > 0.1, a fixed-effects meta-analysis was conducted. We excluded cases with missing values instead of any imputation when we fitted the data for each outcome. To reduce the bias due to the small sample size, we excluded countries with fewer than 25 respondents with disabilities when we pooled the country-specific estimations.

### Ethical Approval

The London School of Hygiene and Tropical Medicine Research Ethics Committee approved this project on the 9th of November 2020 (reference number 22719). Consent was obtained by MICS interviewers at the time of the survey and only participants who consent to have their data shared anonymously are made publicly available on the MICS website. We accessed the anonymized data from the MICS website in January 2023.

## Results

### Overall Sample

Our sample included 513,252 people across 37 countries, with sample sizes ranging from 1,031 in Tuvalu to 57,585 in Bangladesh (Table 2). The overall prevalence of disability in the sample was 4.8% (n = 24,695), but the prevalence ranged from 0.8% of the sample in Turkmenistan (n=58) to 10.8% in Central African Republic (n=1,235) and Costa Rica (n=743). The overall sample was predominantly female (82.5%, n=423,615) and there were a slightly larger proportion of rural participants (55.6%, 285,454).

**Table 2:** Summar statistics of adults 15-49 included in the analysis overall and by country.

| Country | Sample Size | Prevalence of Disability, n (%) | Urban, n (%) | Age, mean (SD) | Male, n (%) | Female, n (%) | Knowledge about HIV, n (%) | Knowledge about MTCT, n (%) | Know of a place to be tested for HIV, n (%) | Ever tested for HIV and know the results, n (%) | Tested for HIV in the last 12 months and know the results, n (%) |
| --- | --- | --- | --- | --- | --- | --- | --- | --- | --- | --- | --- |
| Total sample | 513,252 | 24,695 (4.8) | 227,798 (44.4) | 31.46 (8.96) | 89,637 (17.5) | 423,615 (82.5) | 98,876 (20.0) | 132,846 (26.7) | 281,168 (56.5) | 175,159 (37.9) | 71,772 (26.0) |
| **East Asia and Pacific** |  |  |  |  |  |  |  |  |  |  |  |
| Fiji | 6,827 | 206 (3.0) | 3,849 (56.4) | 33.16 (8.87) | 2,261 (33.1) | 4,566 (66.9) | 2,527 (37.0) | 1,326 (19.4) | 4,861 (71.3) | 2,068 (30.5) | 439 (16.4) |
| Kiribati | 5,669 | 297 (5.2) | 2,402 (42.4) | 31.24 (8.86) | 1,865 (32.9) | 3,804 (67.1) | 2,296 (40.5) | 1,709 (30.2) | 4,672 (82.4) | 557 (9.8) | 220 (9.7) |
| Mongolia | 13,898 | 1,198 (8.6) | 7,118 (51.2) | 34.31 (8.41) | 4,032 (29.0) | 9,866 (71.0) | § | 742 (5.3) | 8,974 (64.6) | 7,042 (50.9) | 3,078 (29.1) |
| Samoa | 4,696 | 77 (1.6) | 1,478 (31.5) | 31.82 (9.49) | 1,047 (22.3) | 3,649 (77.7) | 409 (8.7) | 1,118 (23.8) | 1,368 (29.1) | 237 (5.1) | 87 (5.6) |
| Tonga | 3,525 | 79 (2.2) | 991 (28.1) | 31.78 (9.37) | 1,047 (29.7) | 2,478 (70.3) | § | 897 (25.5) | 2,030 (57.6) | 394 (11.2) | 136 (11.7) |
| Tuvalu | 1,031 | 56 (5.4) | 663 (64.3) | 30.56 (8.39) | 270 (26.2) | 761 (73.8) | 248 (24.1) | 233 (22.6) | 669 (65.0) | 294 (28.6) | 82 (13.4) |
| **East and Central Asia** |  |  |  |  |  |  |  |  |  |  |  |
| Belarus | 7,428 | 78 (1.1) | 5,516 (74.3) | 33.91 (7.66) | 2,167 (29.2) | 5,261 (70.8) | 4,103 (55.3) | 1,541 (20.8) | 7,313 (98.5) | 6,592 (89.0) | 2,440 (36.5) |
| Georgia | 8,894 | 807 (9.1) | 4,254 (47.8) | 34.47 (8.61) | 2,467 (27.7) | 6,427 (72.3) | 1,220 (13.7) | 1,859 (20.9) | 3,974 (44.7) | 2,031 (22.9) | 651 (22.2) |
| Kosovo | 8,400 | 587 (7.0) | 3,910 (46.5) | 32.49 (9.52) | 2,383 (28.4) | 6,017 (71.6) | 768 (9.1) | 1,264 (15.1) | § | § | § |
| Kyrgyzstan | 5,162 | 134 (2.6) | 2,355 (45.6) | 33.07 (8.90) | § | 5,162 (100.0) | § | 2,434 (47.2) | 4,506 (87.3) | 3,929 (76.6) | 1,104 (25.5) |
| Montenegro | 2,848 | 36 (1.3) | 1,720 (60.4) | 33.90 (8.57) | 751 (26.4) | 2,097 (73.6) | 918 (32.3) | 692 (24.3) | 1,731 (60.8) | 191 (6.7) | 37 (10.7) |
| Turkmenistan | 6,959 | 58 (0.8) | 3,631 (52.2) | 32.21 (8.74) | § | 6,959 (100.0) | 2,166 (31.1) | 1,709 (24.6) | 4,546 (65.4) | 1,996 (29.1) | 677 (14.6) |
| Uzbekistan | 4,385 | 289 (6.6) | 2,146 (48.9) | 32.76 (8.63) | § | 4,385 (100.0) | 726 (16.6) | 745 (17.0) | 2,800 (63.9) | 2,062 (47.6) | 807 (27.5) |
| **Latin America and the Caribbean** |  |  |  |  |  |  |  |  |  |  |  |
| Costa Rica | 6,894 | 743 (10.8) | 4,316 (62.6) | 31.99 (8.63) | § | 6,894 (100.0) | 2,098 (30.4) | 1,250 (18.1) | 5,743 (83.3) | 4,234 (61.8) | 879 (19.1) |
| Cuba | 11,847 | 151 (1.3) | 8,392 (70.8) | 33.17 (8.76) | 3,453 (29.1) | 8,394 (70.9) | 6,708 (56.7) | 4,213 (35.6) | 11,304 (95.4) | 10,014 (84.9) | 3,510 (32.8) |
| Dominican Republic | 19,992 | 803 (4.0) | 13,905 (69.6) | 32.11 (9.16) | § | 19,992 (100.0) | 7,616 (38.1) | 8,846 (44.3) | 19,106 (95.6) | 16,778 (84.1) | 6,361 (36.7) |
| Guyana | 7,209 | 233 (3.2) | 2,016 (28.0) | 31.77 (9.31) | 1,962 (27.2) | 5,247 (72.8) | 2,679 (37.2) | 2,419 (33.6) | 6,465 (89.8) | 5,021 (70.0) | 1,973 (35.9) |
| Honduras | 23,893 | 1,983 (8.3) | 9,680 (40.5) | 31.28 (9.14) | 6,776 (28.4) | 17,117 (71.6) | 4,861 (20.3) | 8,217 (34.4) | 17,272 (72.3) | 11,733 (49.3) | 3,008 (21.3) |
| Suriname | 8,649 | 459 (5.3) | 5,966 (69.0) | 32.25 (9.01) | 2,420 (28.0) | 6,229 (72.0) | 3,327 (38.5) | 2,926 (33.9) | 7,357 (85.1) | 5,454 (63.3) | 1,984 (31.9) |
| **Middle East and North Africa** |  |  |  |  |  |  |  |  |  |  |  |
| Algeria | 31,368 | 1,444 (4.6) | 21,152 (67.4) | 32.62 (8.95) | § | 31,368 (100.0) | § | 5,255 (16.8) | 8,037 (25.7) | 3,035 (9.7) | 915 (8.7) |
| Iraq | 26,724 | 1,338 (5.0) | 17,912 (67.0) | 31.20 (9.07) | § | 26,724 (100.0) | 1,314 (4.9) | 2,257 (8.4) | 3,854 (14.4) | 1,218 (4.6) | 358 (2.5) |
| Palestine | 9,777 | 221 (2.3) | 5,800 (59.3) | 30.73 (8.93) | § | 9,777 (100.0) | 631 (6.5) | 1,286 (13.2) | § | § | § |
| Tunisia | 11,993 | 908 (7.6) | 7,949 (66.3) | 33.31 (9.04) | 2,232 (18.6) | 9,761 (81.4) | 1,695 (14.1) | 1,989 (16.6) | 3,421 (28.5) | 336 (2.8) | 96 (4.6) |
| **South Asia** |  |  |  |  |  |  |  |  |  |  |  |
| Bangladesh | 57,585 | 1,773 (3.1) | 11,791 (20.5) | 31.59 (8.87) | § | 57,585 (100.0) | 5,067 (8.8) | 5,912 (10.3) | 9,443 (16.4) | § | § |
| Nepal | 18,166 | 362 (2.0) | 10,716 (59.0) | 31.04 (8.74) | 4,853 (26.7) | 13,313 (73.3) | 3,781 (20.8) | 2,371 (13.1) | 10,467 (57.6) | 2,676 (14.8) | 640 (9.5) |
| **Sub-Saharan Africa** |  |  |  |  |  |  |  |  |  |  |  |
| Central African Republic | 11,462 | 1,235 (10.8) | 5,052 (44.1) | 30.05 (8.64) | 3,357 (29.3) | 8,105 (70.7) | 1,717 (15.0) | 3,880 (33.9) | 6,761 (59.0) | 4,721 (41.3) | 2,143 (28.5) |
| Chad | 24,931 | 1,294 (5.2) | 5,502 (22.1) | 29.96 (8.66) | 5,661 (22.7) | 19,270 (77.3) | 4,783 (19.2) | 8,387 (39.8) | 13,265 (53.3) | 6,244 (25.2) | 2,771 (25.5) |
| Democratic Republic of Congo | 24,160 | 953 (3.9) | 7,836 (32.4) | 30.17 (8.70) | 5,187 (21.5) | 18,973 (78.5) | 4,209 (17.4) | 4,971 (20.6) | 11,455 (47.4) | 5,033 (20.9) | 2,122 (22.5) |
| Gambia | 15,520 | 470 (3.0) | 7,929 (51.1) | 30.06 (8.53) | 3,743 (24.1) | 11,777 (75.9) | 3,678 (23.7) | 6,502 (41.9) | 11,228 (72.4) | 6,439 (41.6) | 2,165 (31.3) |
| Ghana | 16,821 | 1,355 (8.1) | 8,111 (48.2) | 31.17 (9.31) | 4,305 (25.6) | 12,516 (74.4) | 2,792 (16.6) | 6,894 (41.0) | 11,862 (70.5) | 6,832 (40.7) | 2,340 (26.7) |
| Guinea Bissau | 11,992 | 252 (2.1) | 4,078 (34.0) | 30.03 (8.62) | 2,391 (19.9) | 9,601 (80.1) | 1,606 (13.4) | 5,022 (41.9) | 6,299 (52.5) | 3,824 (31.9) | 1,256 (23.4) |
| Madagascar | 21,333 | 1,608 (7.5) | 6,029 (28.3) | 30.27 (9.08) | 6,467 (30.3) | 14,866 (69.7) | 4,754 (22.3) | 4,459 (20.9) | 7,236 (33.9) | 2,512 (11.8) | 665 (6.9) |
| Malawi | 26,724 | 1,523 (5.7) | 4,428 (16.6) | 30.21 (8.89) | 5,610 (21.0) | 21,114 (79.0) | 11,366 (42.6) | 16,159 (60.5) | 26,236 (98.2) | 24,555 (91.9) | 15,565 (62.3) |
| Sao Tome and Principe | 3,794 | 268 (7.1) | 2,177 (57.4) | 31.23 (8.98) | 1,159 (30.5) | 2,635 (69.5) | 1,448 (38.2) | 914 (24.1) | 3,560 (93.9) | 3,043 (80.7) | 1,651 (50.9) |
| Sierra Leone | 21,999 | 286 (1.3) | 8,678 (39.4) | 30.33 (8.75) | 6,371 (29.0) | 15,628 (71.0) | 5,298 (24.1) | 8,458 (38.5) | 14,509 (66.0) | 8,060 (36.8) | 2,262 (17.0) |
| Togo | 8,367 | 629 (7.5) | 3,478 (41.6) | 31.37 (8.74) | 1,959 (23.4) | 6,408 (76.6) | 2,067 (24.7) | 3,990 (47.7) | 6,719 (80.3) | 4,925 (58.9) | 1,730 (30.6) |
| Zimbabwe | 12,330 | 502 (4.1) | 4,872 (39.5) | 31.27 (8.96) | 3,441 (27.9) | 8,889 (72.1) | § | § | 12,125 (98.3) | 11,079 (89.9) | 7,620 (67.7) |

§ = country did not have data available

### Comprehensive knowledge about HIV prevention

Thirty-two countries reported data on comprehensive knowledge about HIV prevention in women (Figure 1). Overall, the pooled showed women with disabilities have substantially lower knowledge about HIV prevention than women without disabilities. For example, no women with disabilities in Samoa had comprehensive knowledge of HIV prevention and, in Sierra Leone, women with disabilities less likely to have comprehensive HIV knowledge than women without disabilities (aRR: 0.33, 95% C.I.: 0.20, 0.57). Belarus, Chad, Democratic Republic of Congo, and Kosovo also had evidence of substantial relative inequities for women with disabilities. In Tuvalu, however, there was some evidence women with disabilities had more knowledge about HIV prevention than women without disabilities (aRR: 1.62, 95% C.I.: 1.05, 2.49).

**Figure 1:**
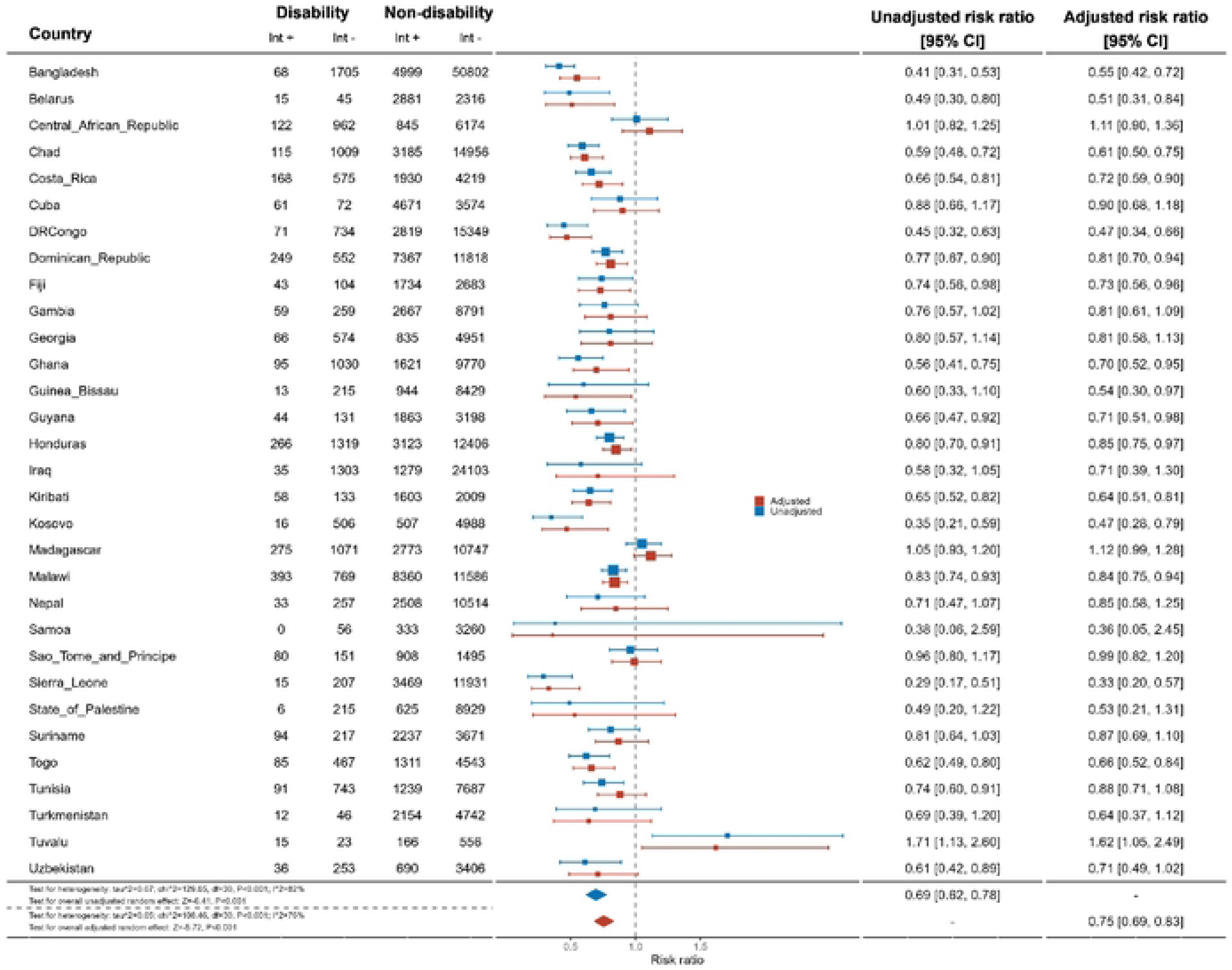
Meta-analysis comparing comprehensive knowledge about HIV prevention among women with disabilities compared to women without disabilities.

Nineteen countries reported data on comprehensive HIV knowledge for men (Figure 2). It showed that men with disabilities were significantly less likely to have comprehensive knowledge of HIV prevention than men without disabilities (aRR: 0.74, 95%C.I.: 0.67, 0.81). This difference was most pronounced in Ghana (aRR: 0.46, 95% C.I.: 0.27, 0.78) and Chad (aRR: 0.53, 95% C.I.: 0.33, 0.84). Most countries had smaller sample sizes and wider confidence intervals for these analyses.

**Figure 2:**
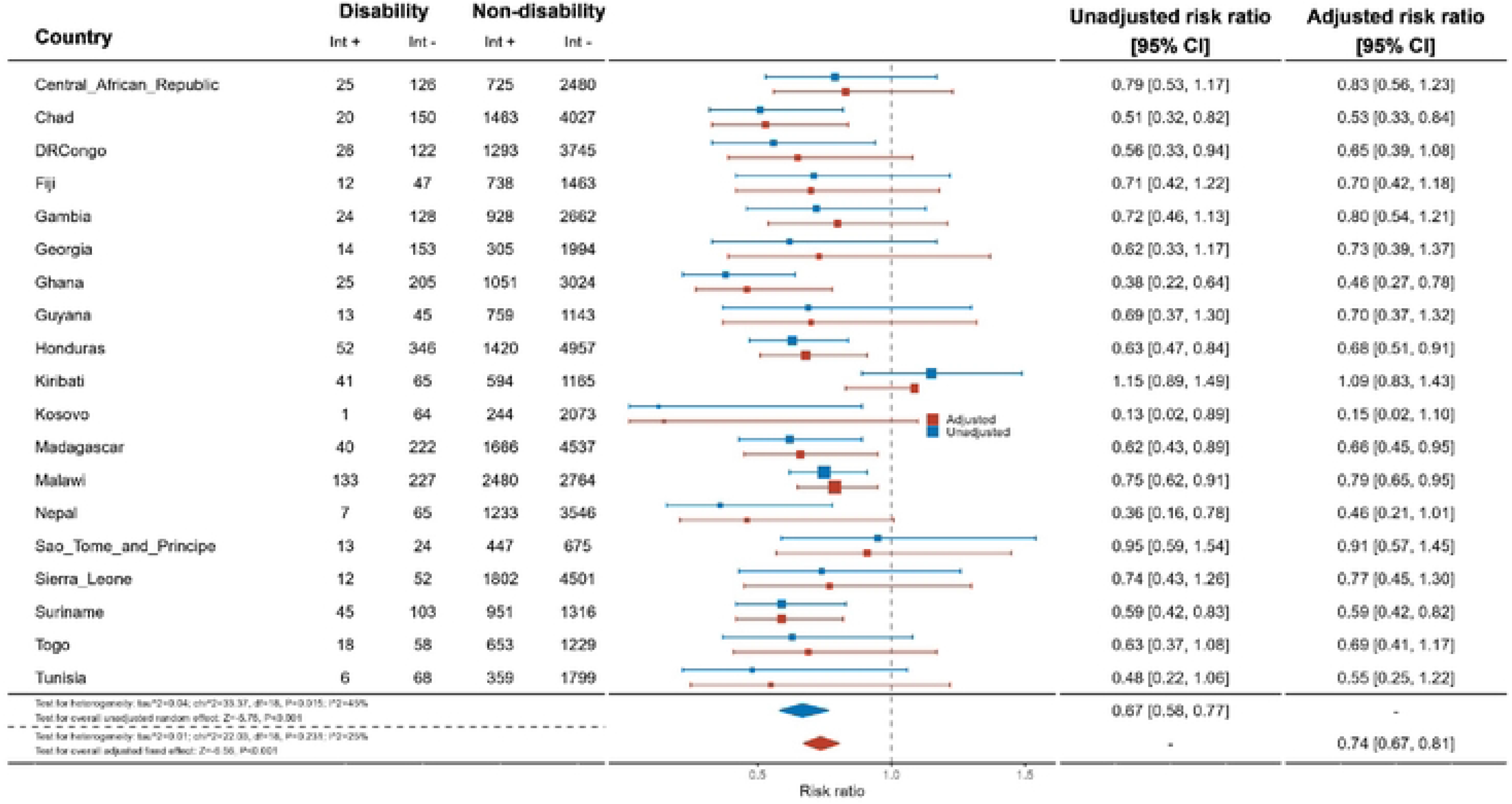
Meta-analysis comparing comprehensive knowledge about HIV prevention among men with disabilities compared to men without disabilities.

### Knowledge of Mother-to-Child Transmission

Women with disabilities were less likely to have knowledge of mother-to-child transmission (MTCT) than women without disabilities for the pooled sample of 37 countries (aRR: 0.87, 95% C.I.: 0.81, 0.93) (Figure 3). Palestine (aRR: 0.39, 95% C.I.: 0.21, 0.72), Kyrgyzstan (aRR: 0.60, 95% C.I. 0.44, 0.82), and Sierra Leone (aRR: 0.59, 95% C.I.: 0.46, 0.77) had the most marked differences between women with and without disabilities, while most other countries had wide confidence intervals and uncertain results. Conversely, women with disabilities were more likely to have knowledge of MTCT in Madagascar (aRR: 1.31, 95%C.I.: 1.16, 1.47) and Sao Tome et Principe (aRR: 1.27, 95%C.I.: 1.00, 1.63).

**Figure 3:**
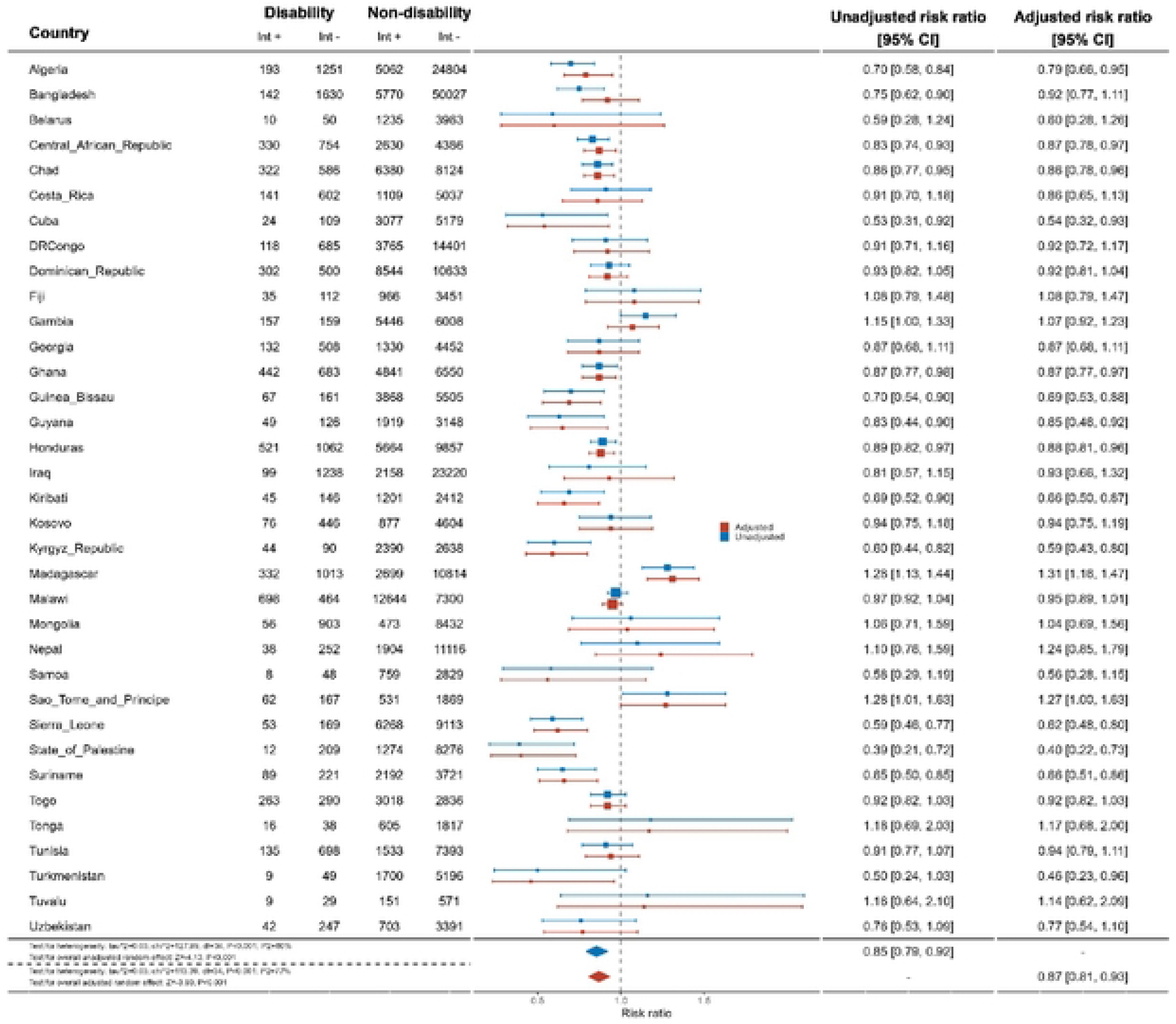
Meta-analysis comparing knowledge of mother-to-child transmission among women with disabilities compared to women without disabilities.

By contrast, across 21 countries, there was no evidence that men with disabilities had less knowledge about MTCT than men without disabilities (Figure 4). Exceptions were Malawi (aRR: 0.77, 95% C.I.: 0.65, 0.93) and Suriname (aRR: 0.62, 95% C.I.: 0.40, 0.96), where men with disabilities were less likely to have MTCT knowledge.

**Figure 4:**
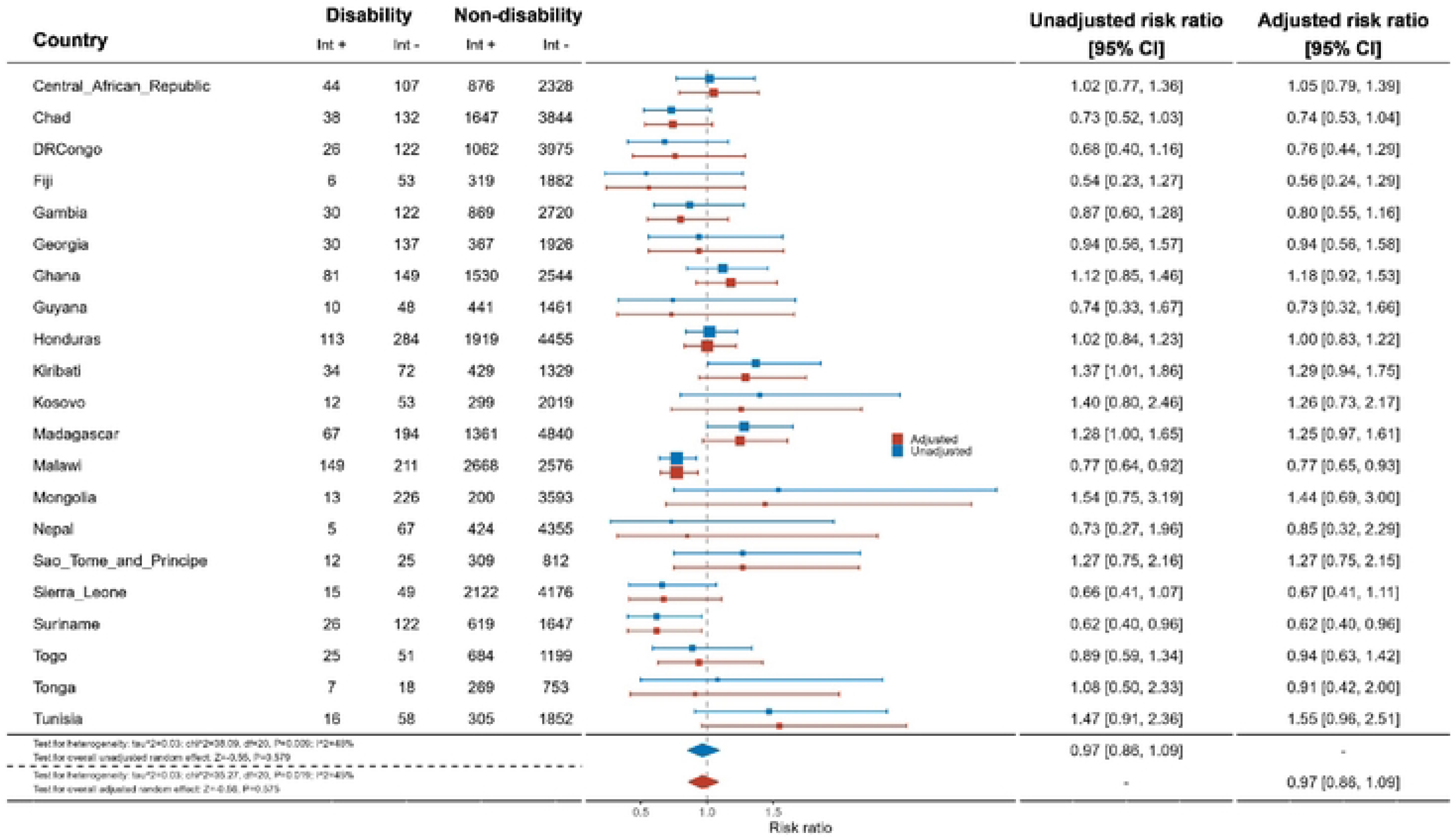
Meta-analysis comparing knowledge of mother-to-child transmission among men with disabilities compared to men without disabilities.

### People who know where to be tested for HIV

Data from 32 countries suggested that women with disabilities were less likely to know where to be tested for HIV than women without disabilities (aRR: 0.94, 95% C.I.: 0.92, 0.97) (Figure 5). This difference was most substantial in Turkmenistan (aRR: 0.59, 95%C.I.: 0.42, 0.82). In contrast, in Tunisia (aRR: 1.23, 95% C.I.:1.10, 1.37) and Madagascar (aRR: 1.12, 95% C.I.: 1.03, 1.21), women with disabilities were slightly more likely to know where to be tested for HIV than women without disabilities.

**Figure 5:**
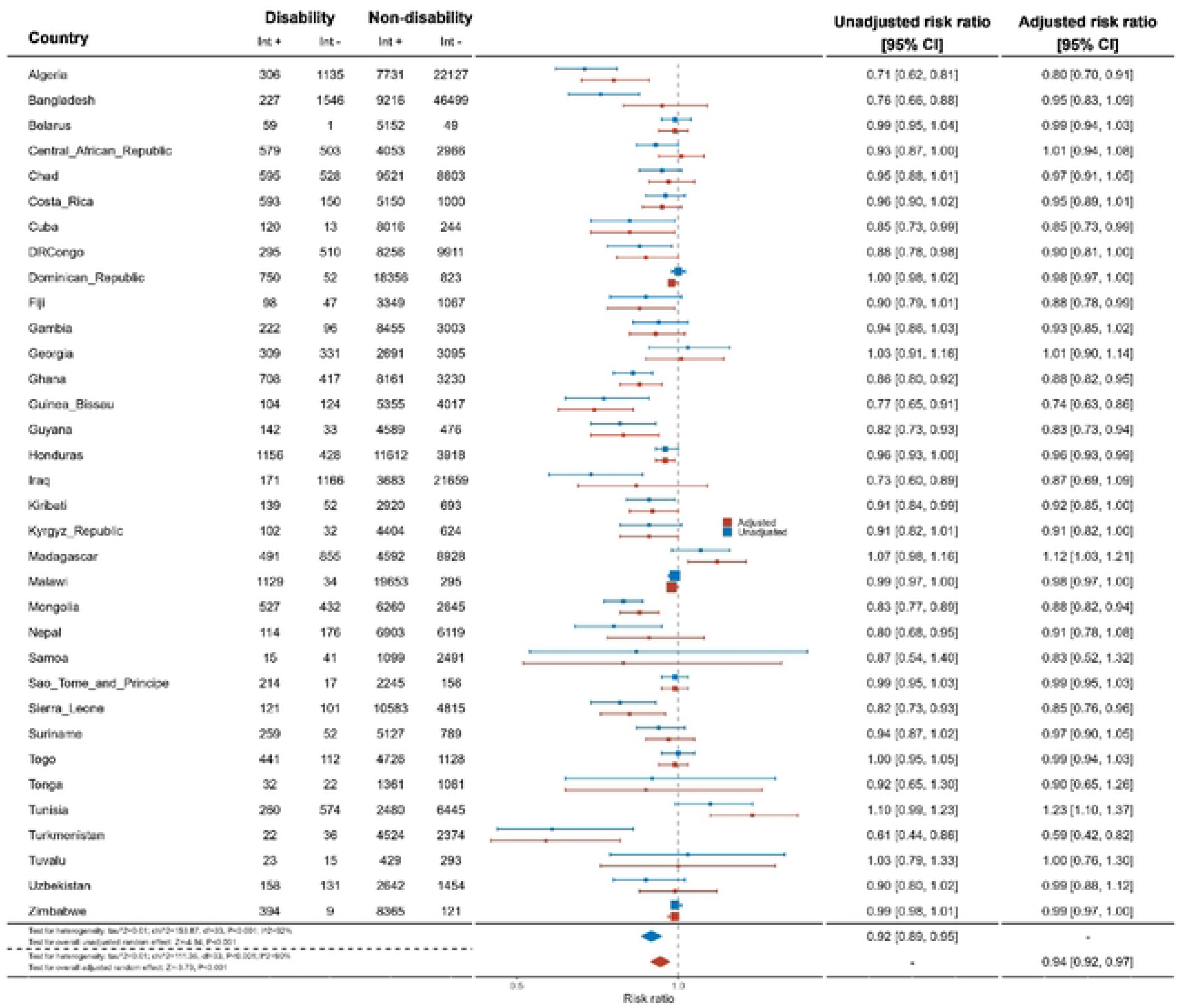
Meta-analysis of women with disabilities who know of a place to be tested for HIV compared to women without disabilities.

Results for men, across 21 countries, also showed men with disabilities were less likely to know where to get tested for HIV than men without disabilities (aRR: 0.95, 95% C.I.: 0.92, 0.99) (Figure 6). However, most individual country results were uncertain.

**Figure 6:**
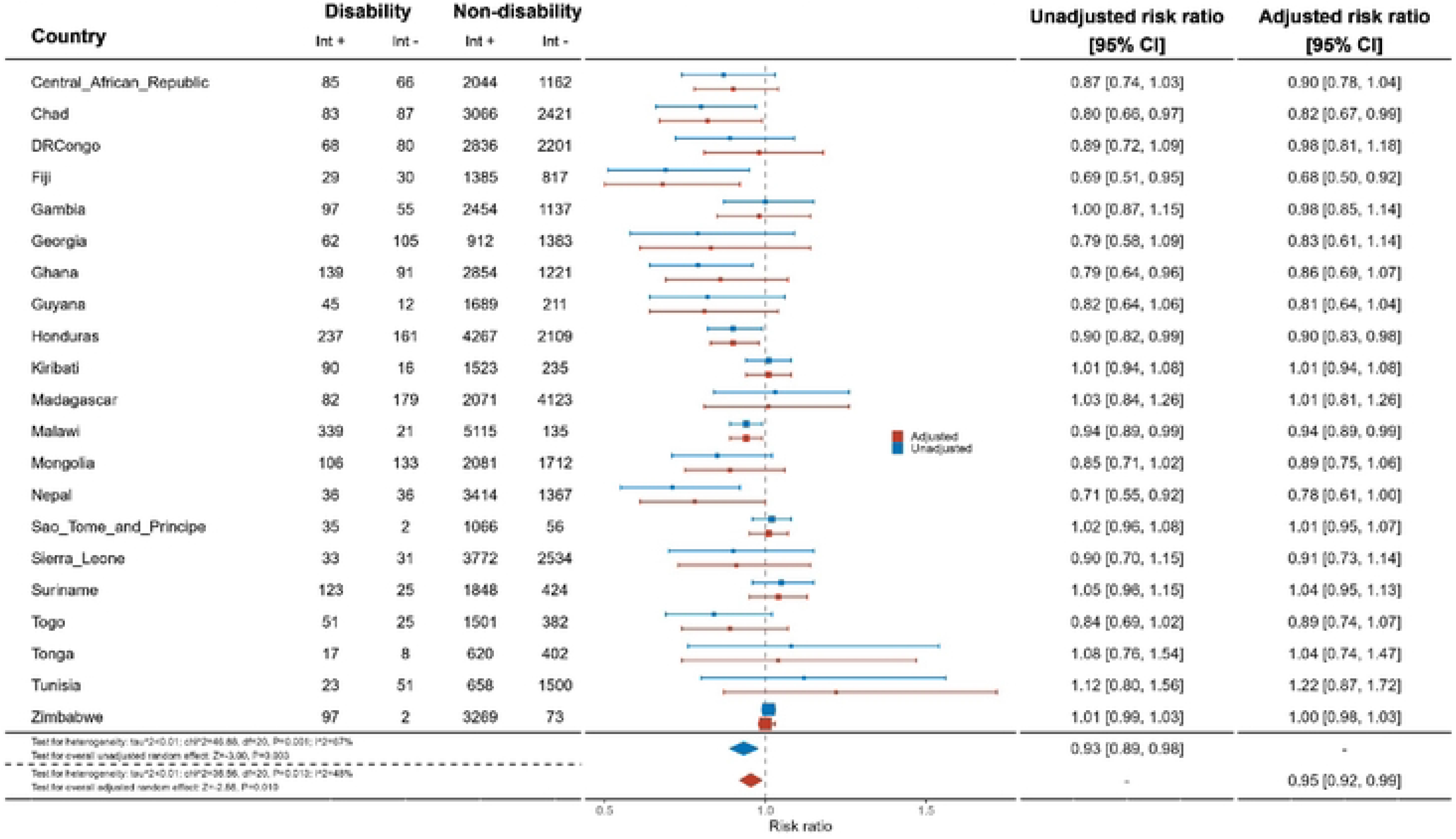
Meta-analysis of men with disabilities who know of a place to be tested for HIV compared to men without disabilities.

### People who have ever been tested for HIV and know the results

Across 32 countries, women with disabilities were less likely to have ever been tested and know their results for HIV than women without disabilities (aRR: 0.90, 95% C.I.: 0.85, 0.94) (Figure 7). This difference was most pronounced in Guinea-Bissau (aRR: 0.62, 95% C.I.: 0.49, 0.79). Most countries showed strong evidence of relative inequities. However, in Tunisia, women with disabilities were more likely to have ever been tested for HIV than women without disabilities (aRR: 1.57, 95% C.I.: 1.07, 2.26).

**Figure 7:**
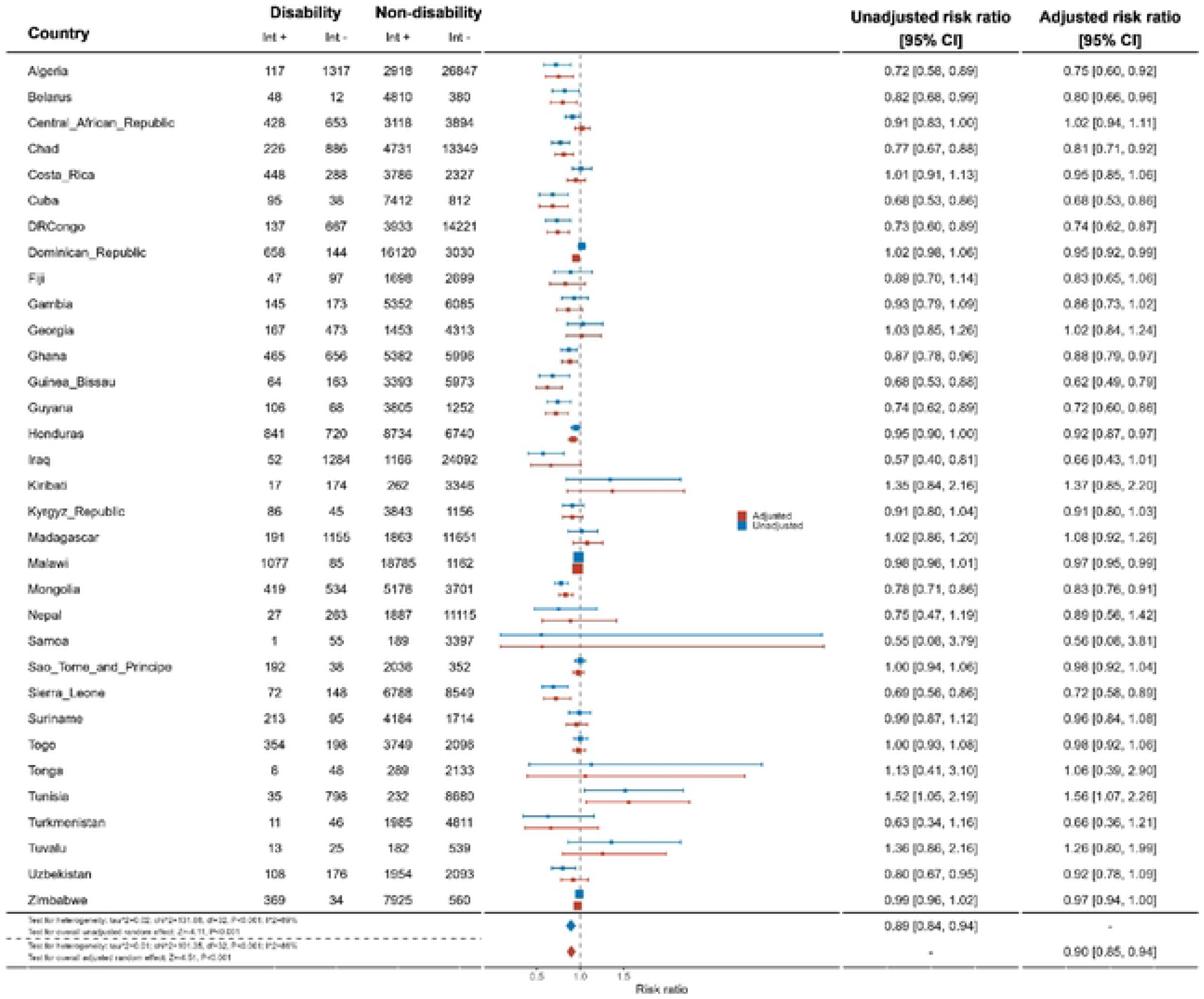
Meta-analysis of women with disabilities who have ever been tested for HIV and know the results compared to women without disabilities.

There was limited evidence that men with disabilities were less likely to have ever been tested for HIV and know their results (aRR: 0.94, 95% C.I.: 0.86, 1.03) (Figure 8). Most countries showed no difference between men with and without disabilities, except in Fiji (aRR: 0.20, 95% C.I.: 0.06, 0.65) and Georgia (aRR: 0.57, 95% C.I.: 0.34, 0.95). As for women, men with disabilities in Tunisia were found to be more likely to have ever been tested for HIV and know their results (aRR: 2.81, 95% C.I.: 1.17, 6.72).

**Figure 8:**
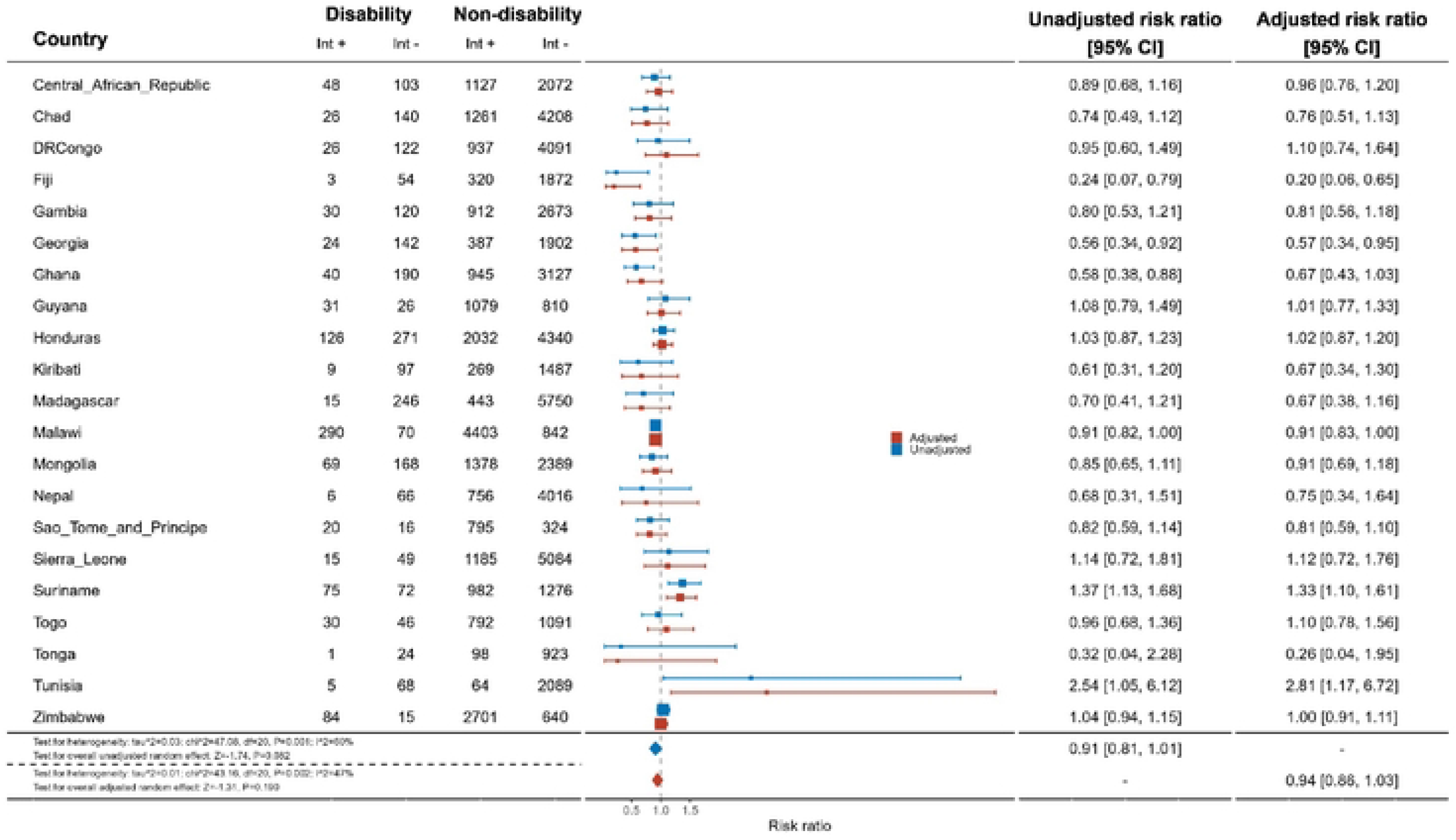
Meta-analysis of men with disabilities who have ever been tested for HIV and know the results compared to men without disabilities.

### People who have been tested for HIV in the past 12 months and know the results

There was some evidence women with disabilities were less likely to have been tested for HIV in the past 12 months and know the results compared to women without disabilities (aRR: 0.95, 95%C.I.: 0.90, 1.02) (Figure 9). There was evidence that women with disabilities were less likely to be tested and know the results in the past 12 months in Algeria (aRR: 0.51, 95% C.I.: 0.32, 0.83), Chad (aRR: 0.72, 95% C.I.: 0.57, 0.90), and Mongolia (aRR: 0.76, 95% C.I.: 0.62, 0.93).

**Figure 9:**
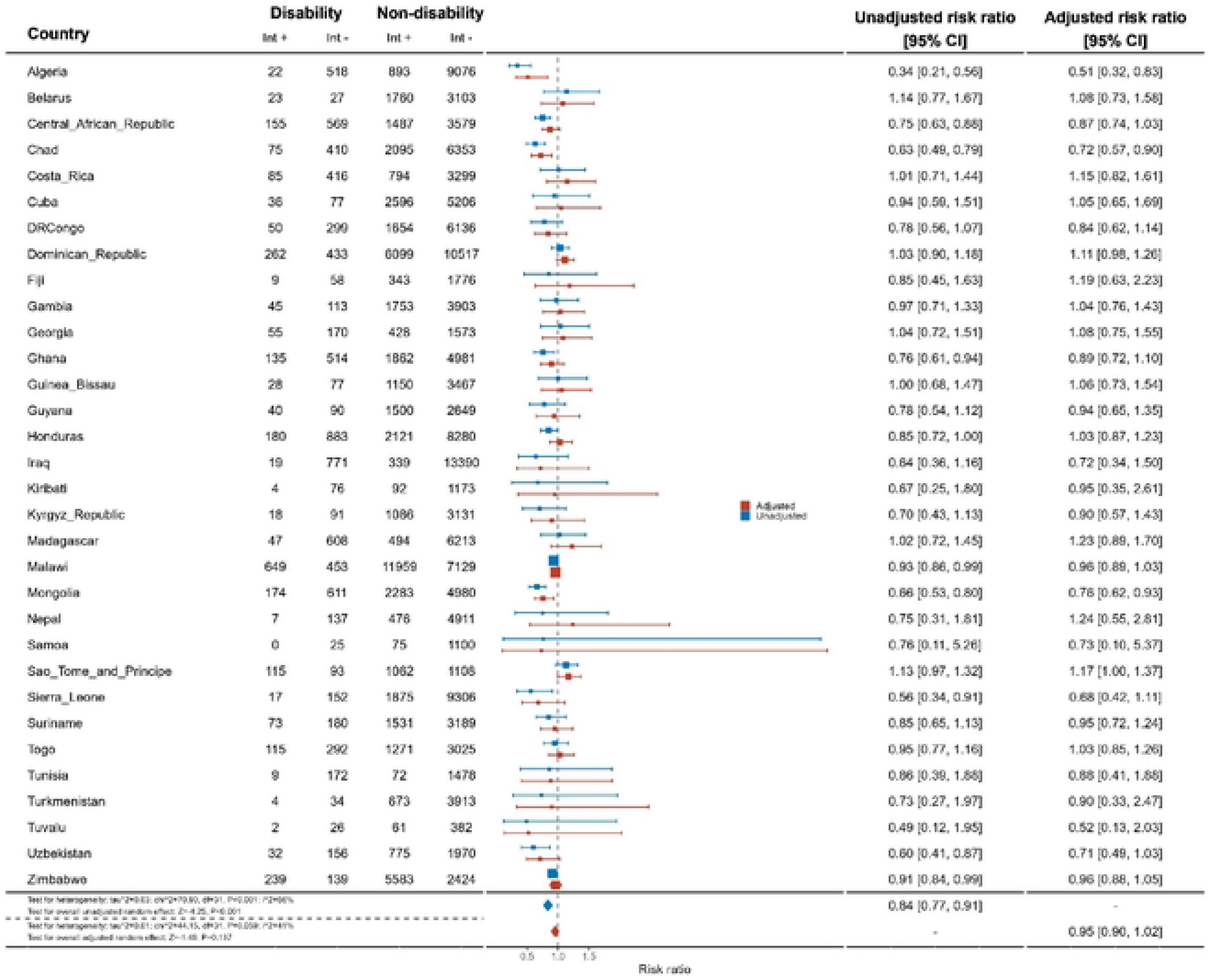
Meta-analysis of women with disabilities who have been tested for HIV in the last 12 months and know the results compared to women without disabilities.

Across 17 countries, men with disabilities were no less likely to have been tested and know the results of the test in the last 12 months than men without disabilities (aRR: 1.02, 95% C.I.: 0.87, 1.20) (Figure 10). This differed in Mongolia, where men with disabilities were less likely to have been tested (aRR: 0.51,.95% C.I.: 0.27, 0.96), while in Suriname (aRR: 1.74, 95% C.I.: 1.28, 2.36) and Togo (aRR: 1.59, 95%C.I.: 1.02, 2.47), men with disabilities were more likely to have been tested in the past 12 months and know the results.

**Figure 10:**
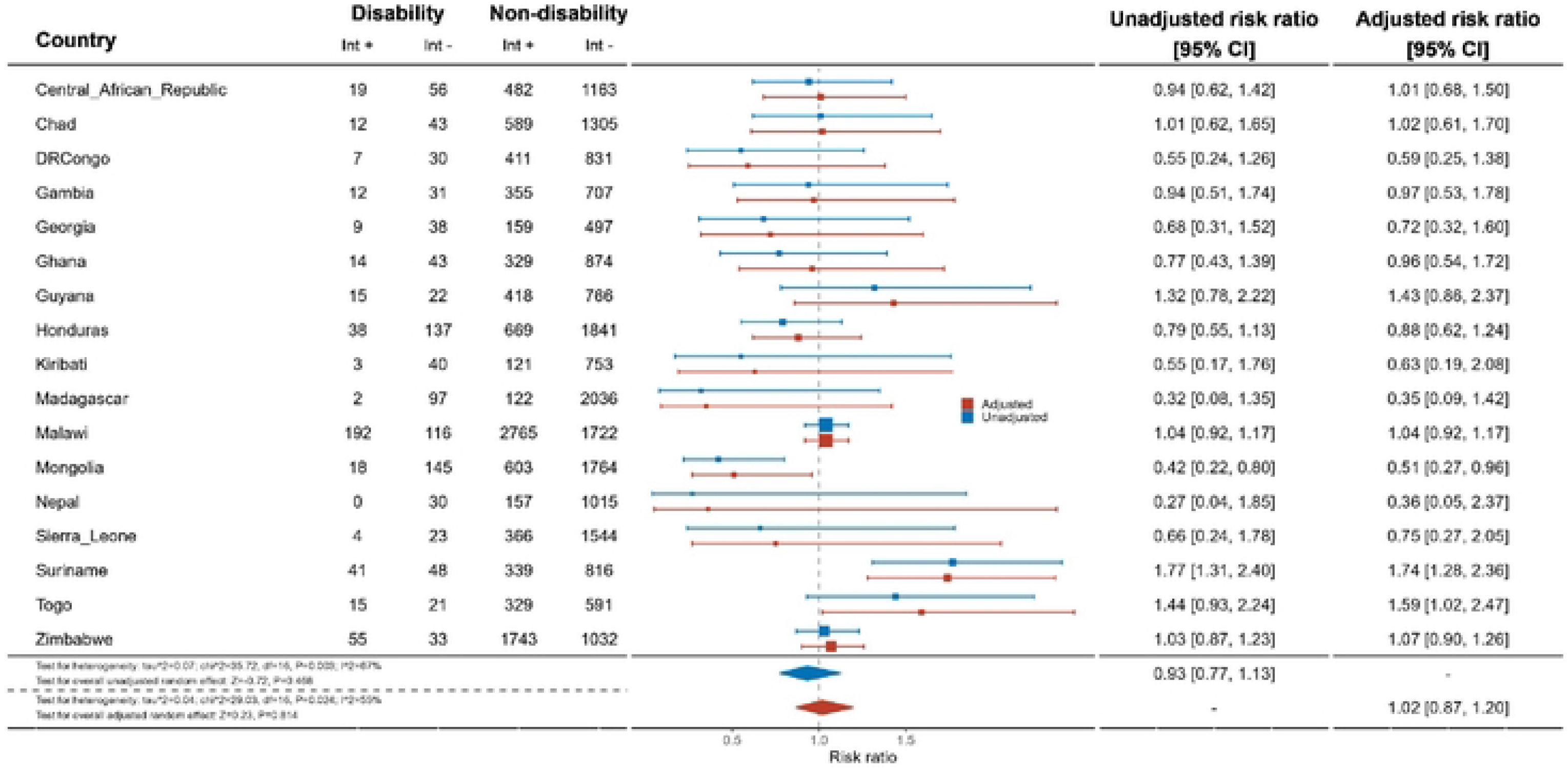
Meta-analysis of men with disabilities who have been tested for HIV in the last 12 months and know the results compared to men without disabilities.

## Discussion

This study provides the largest published evidence on the HIV knowledge and testing gap for people with disabilities across 37 countries. Our findings suggest women with disabilities were less likely to have comprehensive knowledge about HIV prevention, knowledge of MTCT, know where to be tested for HIV compared to women without disabilities, and have ever been tested for HIV and know the results. Men with disabilities were less likely to have comprehensive knowledge about HIV prevention and know of a place to be tested. There was limited evidence that men with disabilities were less likely to have ever been tested for HIV and that women with disabilites had been tested for HIV in the past 12 months and know the results. By contrast, our overall estimate found no differences in having been tested for HIV in the past 12 months and know the result and knowledge of MTCT for men. However, this estimate varied substantially by country and was impacted by small sample sizes, which may explain the result, rather than improved knowledge or access to testing.

These findings are largely consistent with the existing literature that highlight the gaps in HIV knowledge and testing for people. For example, these results are similar to studies in South Africa that showed people with disabilities have less knowledge about HIV and testing sites,[15] as well DHS data from Uganda that showed gaps in transmission.[16] However, the most important finding of this analysis is the gap between men and women with disabilities and showcases the ‘double disadvantage’ women with disabilities experience on the basis of gender and disability. Across all five indicators, women had at least some evidence they were less likely to have knowledge about HIV and access to testing than women without disabilities, whereas this was only the case for two indicators for men. Importantly, this difference was most pronounced for knowledge about MTCT, since there was significantly less knowledge among women with disabilities compared to women without, but no differences among men with and without disabilities. This information is important for all women of childbearing age, but particularly populations where there is a higher prevalence of HIV, including among those with disabilities. This knowledge gap will not only hamper women with disabilities ability to prevent MTCT among their children, but also make the global goal of eliminating MTCT impossible.

These findings are important as they reinforce the concern that Global AIDS targets will not be met without more efforts to include people with disabilities in HIV programmes.[3] Efforts are therefore needed to reach people with disabilities who may be left behind in existing HIV programmes. Health systems can address these gaps by going beyond mentioning disability in their HIV policies, and, instead, integrating specific considerations into their programs and national plans,[4] and across each building block of the health system. To develop these plans fully, governments should look at their leadership, governance, and financing structures to ensure people with disabilities involved in the development of HIV plans and specific budget item lines to address disability inclusion. Efforts should focus on improving the accessibility of HIV services for people with disabilities through ensuring the physical access of the facility. Training health workers about disability, including destigmatizing disability and sexual activity and communicating public health information about HIV as well as individual patient communications are in accessible formats can further improve the quality of these services.

Finally, there needs to be more and comparable data on disability within routine HIV surveys as well as other national and household surveys that look at HIV prevalence, knowledge, and testing. Routine data on the prevalence of HIV among this population will help to monitor efforts to close gaps, as well as further elucidate the relationship between HIV and disability, and disaggregate further by other vulnerabilities (e.g., education, violence, and social isolation). Indeed analysis of the Demographic Health Survey in South Africa shows that women with disabilities, who were also living with HIV were 4 times as likely to experience intimate partner violence then those without disabilities and HIV.[24] Together, these efforts will improve knowledge and testing among people with disabilities and so close the gaps. Good practice examples exist already of disability-inclusion in HIV services, such as in Jamacia where HIV-focused civil society organizations are collaborating with organizations of persons with disabilities to reach people with disabilities,[25] and South Africa, where health workers are being trained about disability and HIV,[26] but these need to be scaled further.

### Strengths and Limitations

This analysis the largest examining HIV knowledge and testing by disability status. It allows cross-country comparison, providing new evidence from countries outside of sub-Saharan Africa, while also furthering the breadth of evidence in disability-based HIV knowledge and testing inequities in this region. Disaggregation of data by sex also allows us to examine the inequities by disability and sex, which revealed greater absolute inequities for women with disabilities. Given the global focus on improving gender-based inequities, this analysis provides important evidence on these gaps and how women with disabilities need to be further included in gender-targeted programs. Combining this information with other studies also calls for more nuanced research to understand which people with disabilities are left behind and how this intersects with gender, age and mitigating factors (poverty, isolation, education and exposure to violence).

However, this analysis was limited by the definition of disability used in the MICS, which results in a lower prevalence than is estimated globally and our analysis which focused only on people with at least a lot of difficulties in one domain. In addition, the Washington Group Short Set used for people aged 18-49 omit the full experience of disability, particularly those with psychosocial, intellectual, neurological, developmental, and upper limb-based disabilities.[27] As these are cross sectional surveys that do not test the onset of functional limitations, we also cannot understand if the individuals identified as having ‘disabilities’ are those with preexisting disabilities or acquired because of HIV treatment. Additionally, since the interview guide recommends only including people who can respond for themselves, it limits the level of functional difficulty captured in the survey. In particular, people with hearing or intellectual impairments may have been excluded. This bias may limit the applicability of our findings to those with only moderate functional limitations, rather than all people with disabilities, particularly those most likely to be excluded from HIV information and testing. Furthermore, the men’s dataset is not only run in fewer countries, but also has a lower response rate (70-80% compared to the women’s 90-95%). This, on top of the possible accessibility barriers to participating, introduces some non-response bias. Finally, we were limited by the covariates we examined, particularly since there was no MICS estimate of HIV prevalence and small sample sizes for some indicators. Since this would impact access to testing and information, this is also a limitation of this analysis.

## Conclusion

Overall, this study provides new data on the inequities in HIV knowledge and testing for people with severe disabilities. Without concerted efforts to reach people with disabilities in HIV programmes we will not be able to achieve the global goals for HIV,[3] and will leave those at most risk behind. With more data revealing these inequities, the gaps for people with disabilities are now well-understood and require urgent action to address.

## Data Availability

Data from UNICEF-supported MICS are publicly available. Our analysis code is also available online.

https://mics.unicef.org/surveys

## Acknowledgements

We thank the participants who participated in MICS surveys and the survey teams in each country.

## Author Contributions

SR conceived the study and wrote the first draft, SC conducted the analysis, JHH, LMB, CD, and HK edited the manuscript and provided crucial feedback on drafts.

